# Drug repositioning candidates identified using in-silico quasi-quantum molecular simulation demonstrate reduced COVID-19 mortality in 1.5M patient records

**DOI:** 10.1101/2021.03.22.21254110

**Authors:** Joy Alamgir, Masanao Yajima, Rosa Ergas, Xinci Chen, Nicholas Hill, Naved Munir, Mohsan Saeed, Ken Gersing, Melissa Haendel, Christopher G Chute, M. Ruhul Abid

## Abstract

**Background:** Drug repositioning is a key component of COVID-19 pandemic response, through identification of existing drugs that can effectively disrupt COVID-19 disease processes, contributing valuable insights into disease pathways. Traditional non *in silico* drug repositioning approaches take substantial time and cost to discover effect and, crucially, to validate repositioned effects.

**Methods:** Using a novel in-silico quasi-quantum molecular simulation platform that analyzes energies and electron densities of both target proteins and candidate interruption compounds on High Performance Computing (HPC), we identified a list of FDA-approved compounds with potential to interrupt specific SARS-CoV-2 proteins. Subsequently we used 1.5M patient records from the National COVID Cohort Collaborative to create matched cohorts to refine our in-silico hits to those candidates that show statistically significant clinical effect.

**Results:** We identified four drugs, Metformin, Triamcinolone, Amoxicillin and Hydrochlorothiazide, that were associated with reduced mortality by 27%, 26%, 26%, and 23%, respectively, in COVID-19 patients.

**Conclusions:** Together, these findings provide support to our hypothesis that in-silico simulation of active compounds against SARS-CoV-2 proteins followed by statistical analysis of electronic health data results in effective therapeutics identification.

## INTRODUCTION

There have been 529,301 US deaths as of March 12, 2021 due to COVID-19.^I1^ The Food and Drug Administration (FDA) has so far approved three COVID-19 vaccines.^I2^ However, a substantial time lag is expected between the start of vaccinations and effective herd immunity.^3^,^4^ Furthermore, vaccine hesitancy is high in the US with 51% to 72% of the population intending to be vaccinated.^5^,^6^,^7^ Additionally, a global race for vaccine acquisition continues.^8^ As 70% of the population must become immune to interrupt this pandemic9, COVID-19-related deaths will continue in the coming months.^4^ Therefore, drug repurposing is urgently needed to reduce COVID-19 mortality ^5^ while providing insight into disease pathways.^10^

In this study, we tested the hypothesis that in-silico quasi-quantum simulation of FDA-approved compounds against SARS-CoV-2 proteins followed by statistical analysis of 1.5M electronic health data can efficiently identify effective drug repositioning candidates.

## METHODS

### In-silico Quasi Quantum Simulation

We used ARIScience’s previously developed quasi-quantum (QQ) molecular simulation platform to disassemble and analyze the energy distribution of 11 SARS-CoV-2 proteins (**Table 1**) against 1,513 known FDA-approved active ingredients. This proprietary method uses electron density approximations, high probability conformations determinations, and multi-dimensional energy searches to determine intermolecular affinities. Using Java framework for highly parallel processing within a supercomputing node, and SLURM to spread load across nodes these proteins were simulated at neutral pH. Top candidates for each targeted protein were loaded into Jupyter^11^ for consolidated candidate interaction energy ranking. The resulting top candidates were chosen for pharmacological prevalence assessment. Next, top candidates were selected for statistically significant clinical effect validation using the National COVID Cohort Collaborative (N3C) repository.^12^ The goal of in-silico simulation was to identify small molecule drugs with strong affinity to SARS-CoV-2 proteins and potential to interrupt or delay viral activity. Control test for the QQ simulation framework used the human nicotine receptor (alpha 4 beta 2) with nicotine as positive control ligand and albuterol as negative control ligand. The resultant interaction energy from the QQ simulation for nicotine (−0.0026) was substantially lower than that of albuterol (energy: −0.0012) in line with the expectation that nicotine interacts substantially with the nicotine receptor, but albuterol does not. The energy units are in 0.0188 Hartrees.

**Table 1:** Functions of 11 targeted SARS-CoV-2 proteins^38^

| Protein | Function | Number of Simulations Performed |
| --- | --- | --- |
| E | Involved in viral assembly and egress. Interacts with M, N, 3a, and 7a. | 931,518 |
| N | Functions in helical ribonucleoprotein formation, viral RNA replication, virion assembly, and immune evasion. Interacts with M and nsp3. | 2,498,292 |
| NSP1 | Inhibits translation of cellular mRNA. Assists viral gene expression and immune evasion. | 456,945 |
| NSP9 | Binds with the viral genome and promotes viral RNA replication. | 717,612 |
| NSP15 | Prevents immune detection by cleaving uridylates from the 5' ends of the negative-sense viral RNA. Loss of nsp15 affects viral replication and pathogenesis. | 3,258,654 |
| NSP16 | Assists in formation of the viral mRNA capping machinery. | 1,066,011 |
| ORF3a | Linked to virulence, ion channel formation, and virus release. Activates NF-kB and NLRP3 inflammasome | 531,948 |
| ORF7a | Viral antagonist of the host restriction factor BST-2/Tetherin. Interacts with S, M, E, and ORF3a. | 152,922 |
| ORF8 | Possesses a signal sequence for ER import. Disrupts IFN-I signaling when exogenously expressed in cells. Downregulates MHC-I. | 549,792 |
| ORF9b | Localizes to mitochondria and suppresses IFN-I responses. | 740,079 |
| Spike | Essential for membrane fusion and host receptor binding. | 1,938,609 |

### Use of De-identified Patient Records

The N3C securely harmonizes Electronic Health Record (EHR) data from 36 medical centers dating from 01-01-2018. The data were securely transferred to a data enclave and harmonized into a single common model.^13^ As of 12-07-2020, N3C contained 26M total patients, 372k COVID+ patients, 1.1B lab results, 401M drug exposures, and 179M procedures. We used date-shifted data; all dates except for age were shifted +/-180 days. Drug exposure timing was calculated from the Earliest RNA-based SARS-CoV-2 Diagnosis (ERSD) for each patient. N3C data were crucial in assessing clinical significance of in-silico findings using actual clinical EHR data. The reason is interaction of a compound to a protein may result in one of (a) no-effect (b) increase or (c) decrease in protein activity. Item (c) is the desired effect.^14^,^15^

### Death Endpoint Definition

We compiled OHDSI death concepts to define our endpoint.^16^ Deaths within 12 weeks after ERSD (excluding deaths via accidents, falls and burns) were classified as COVID-19 associated.

### Statistical Validation of Effect against Death Endpoint

We performed cohort matching to account for the potential confounder bias in the compared cohorts.^17^,^18^ The 18 pre-COVID-19 diagnosis predictors we used to construct a propensity score model for cohort matching were: gender, age, race, geographical region, Charlson Comorbidity Index (CCI) categories (0, 1, 2-3, 4-5, 6+), prior medical disposition (smoker, diabetic, chronic respiratory disorder, hypertensive), prior access to medical care (through prior monthly outpatient, inpatient and ER visits, medication, procedure rates), BMI, data provider and COVID-19-related dexamethasone use. We used Bayesian logistic model19 to have numerically stable estimates. The propensity scores were used for nearest neighbor matching with replacement **(Figure 1**).^17^ To assess the treatment effect on the treated for the treatment of interest, we fit Bayesian logistic regression model with weighting to account for repeated sampling of control patients due to matching with replacement at 95% credible interval. We developed Java/Python/R routines to prepare and analyze N3C data respectively. With a sample size of 2,318 patients in each matched cohort, we can detect a difference of 1% increase in death between the two groups with 80% power.

**Figure 1:**
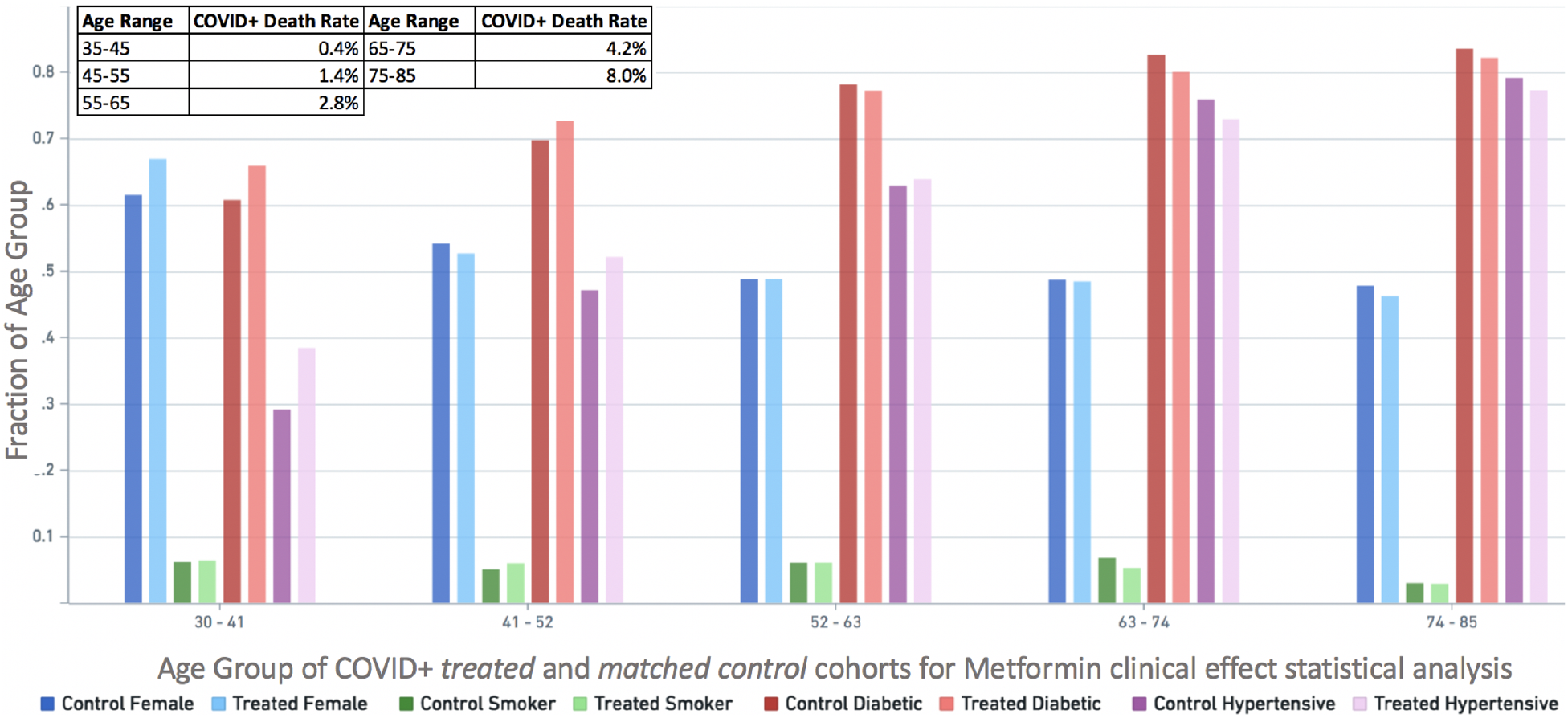
Gender, Smoker, Diabetes, Hypertension by age group (as fraction of age group) for treated (with Metformin) and matched control cohorts across all CCI used for statistical analysis. The darker shade of each colored pair is Control (untreated), while the lighter shade is Treated (with Metformin). For example, the dark purple (control) and light purple (treated) bars on each age group represent the fraction of patients in those age groups that are hypertensive. **Top left box:** death rate by age group for COVID+ patients in N3C.

### Epidemiological Exclusions and Missing Data Handling

To minimize misclassification due to missing data, data-providers were excluded from the analysis if their data was deemed incomplete for key patient information.^20^ Of the 36 data-providers, four were excluded after assessing (a) usage frequency of common medications (Azithromycin, Metformin, Montelukast) (b) medicated patient percentage. This excluded 161,682 patients. We excluded two data-providers whose data quality was not assessed by us affecting 290,578 patients. We excluded six data-providers due to missing death data, excluding 632,614 patients. The final cohort included 1.52M patients.

## RESULTS

### Drug repositioning candidates by in-silico quasi-quantum simulation

The eleven SARS-CoV-2 proteins we chose for computational analyses are: nsp1, nsp9, nsp15, S, N, E, ORF3a, ORF7a, ORF8, and ORF9b (**Table 1**). While nsp1, nsp9, and nsp15 are essential components of viral replication, S, N, and E are the structural proteins needed for production of mature virions. ORF3a, ORF7a, ORF8, and ORF9b are virulence factors that enable the virus to create a favorable replication environment.^21,22,23,24^ After a pharmacological prevalence assessment of the top in-silico candidates and their affinity energies, 18 candidate compounds (**Table 2**) were selected for statistical validation using 1.5M patients.

**Table 2:** Top candidates in ascending order of energy from in-silico simulations followed by pharmacological prevalence assessment. The energy units are in 0.0188 Hartrees.

| Protein | Candidate Drug | Interaction Energy |
| --- | --- | --- |
| N | Metformin | -0.007279 |
| NSP16 | Clindamycin Phosphate | -0.006267 |
| NSP15 | Cromolyn | -0.006233 |
| S | Hydrochlorothiazide | -0.005759 |
| NSP1 | Hydrochlorothiazide | -0.005389 |
| NSP9 | Olmesartan | -0.004378 |
| NSP16 | Bictegravir | -0.003759 |
| NSP1 | Cetirizine | -0.003568 |
| NSP1 | Losartan | -0.003554 |
| NSP1 | Amoxicillin | -0.003459 |
| ORF7a | Metformin | -0.003090 |
| N | Amoxicillin | -0.002472 |
| NSP15 | Vitamin K | -0.002361 |
| NSP16 | Timolol | -0.001635 |
| S | Triamcinolone | -0.001629 |
| NSP9 | Propafenone | -0.001498 |
| NSP16 | Prednisone | -0.001345 |
| NSP16 | Vitamin E | -0.001330 |
| N | Triamcinolone | -0.001145 |
| E | Sildenafil | -0.001008 |
| NSP15 | Simvastatin | -0.000958 |
| S | Fluticasone Furoate | -0.000528 |
| NSP16 | Olmesartan Medoxomil | -0.000324 |

### Clinical effect validation using 1.5M patients’ records

The primary measured endpoint (EP) was death within 84 days of ERSD among 30-to-85 years old (yo) patients. This age range was chosen based upon mortality frequency by age of COVID+ patients in literature (**Figure 1**).^25^ Candidate drugs identified using our in-silico simulation were used to create multi-predictor-based matched cohorts to measure COVID-19 mortality statistical significance. The patients were stratified into three sets with matched cohorts created for each set for each assessed drug. These were: (a) all 30-85yo patients regardless of CCI^26^, (b) all 30-85yo diabetic patients with CCI<=3, and (c) all 30-85yo non-diabetic patients with CCI<=3. The results for each group are as follows:

#### All 30-85yo patients

Metformin, Triamcinolone, Amoxicillin, and Hydrochlorothiazide showed a reduction in mortality odds by 27%, 26%, 26%, and 23%, respectively (**Table 3**). Exposure to these drugs was based on whether they are generally taken chronically (Metformin, Hydrochlorothiazide, non-topical Triamcinolone) or acutely (Amoxicillin). Chronic and acute exposure to a drug considered (a) exposure 365 days prior to ERSD to 2 weeks after ERSD and (b) exposure 4 weeks prior to ERSD to 2 weeks after ERSD, respectively.

**Table 3:**
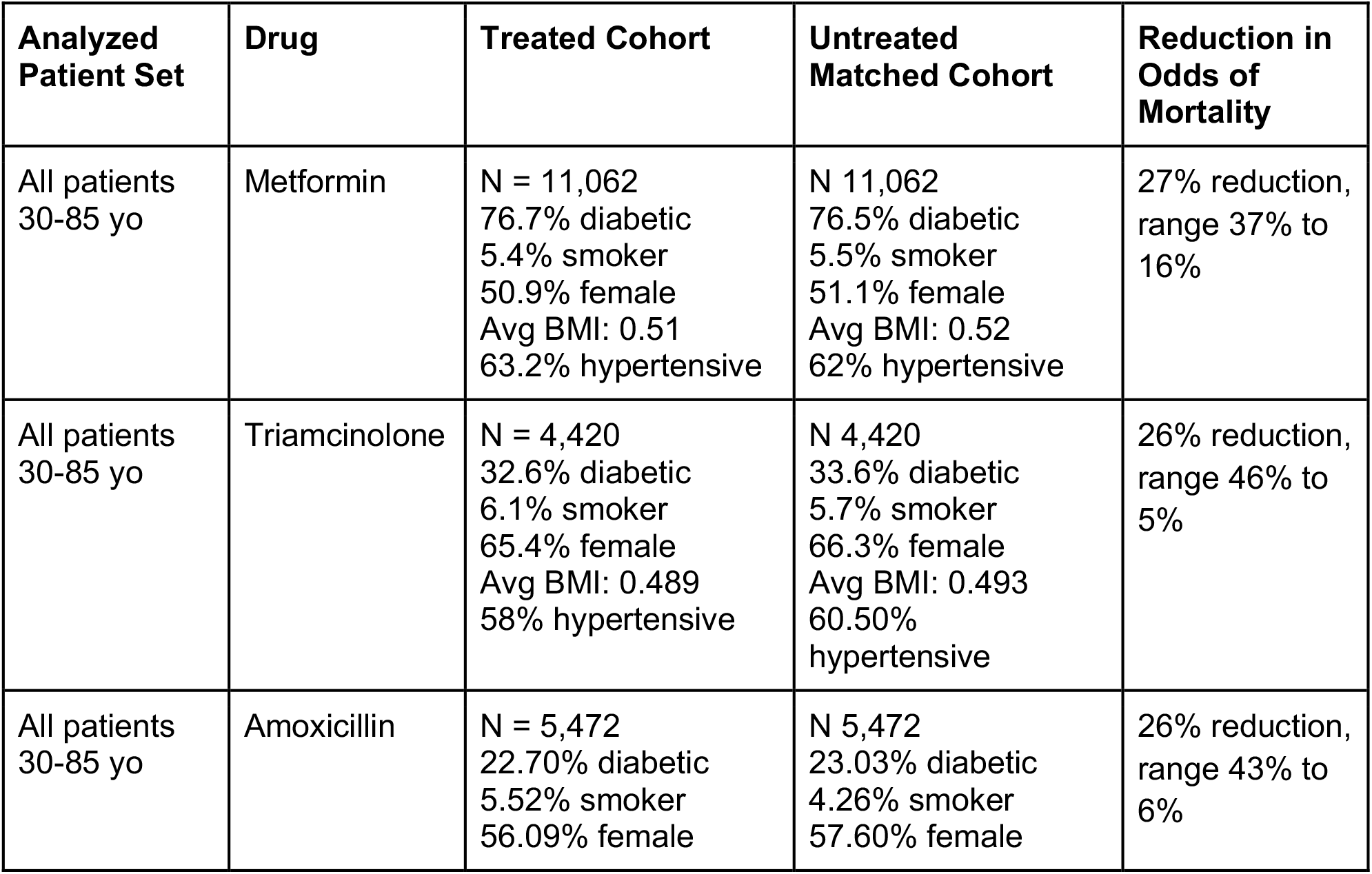
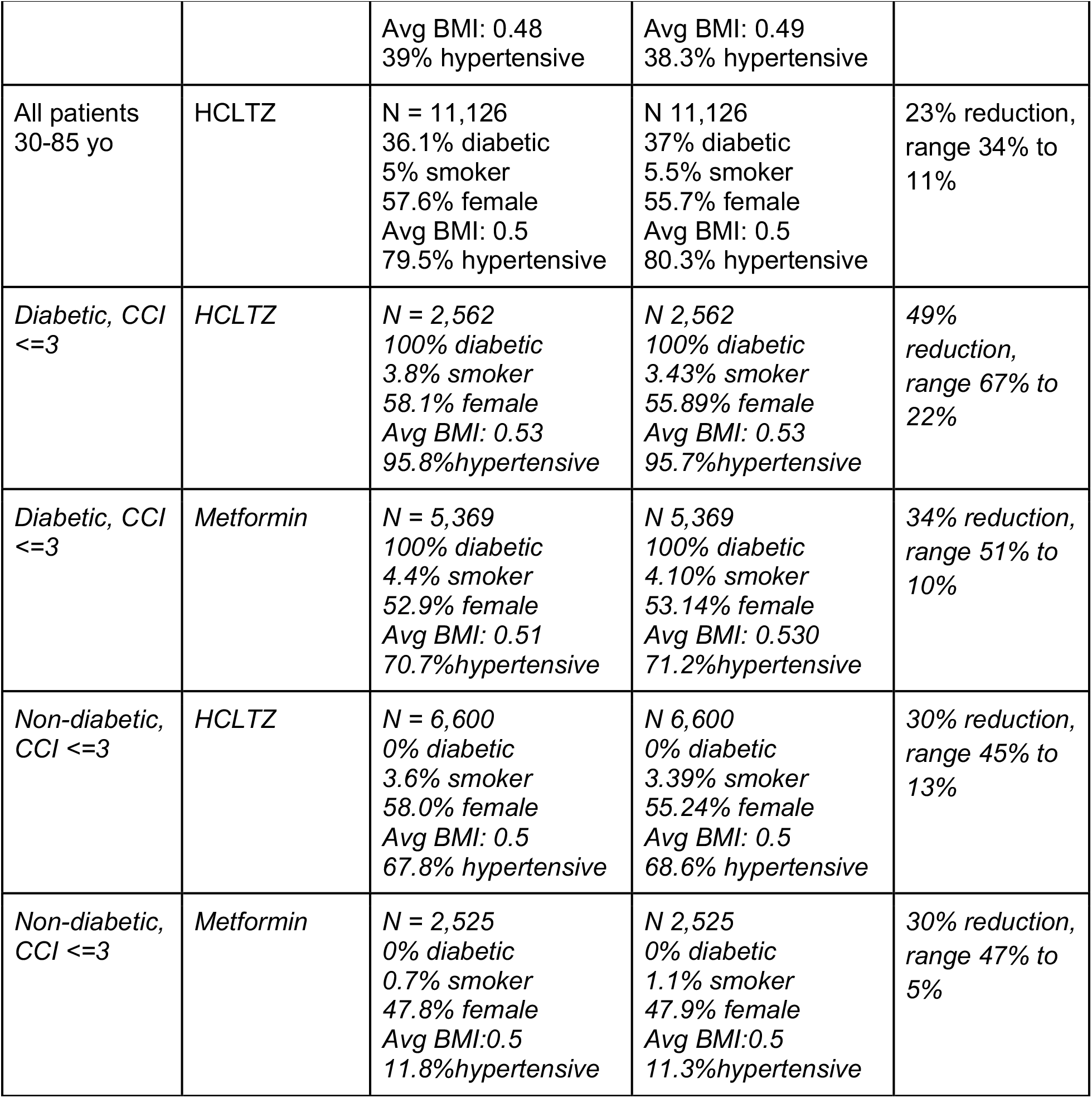
Effect of drug repositioning candidates on mortality odds. HCLTZ is hydrochlorothiazide. BMI scale was modified to be between 0 and 1. Losartan showed statistical significance for the diabetic group but not for the non-diabetic group, and therefore was not included in the above table.

Our in-silico simulations showed Metformin’s affinity to N and ORF7a proteins with energies of − 0.0072 and −0.00309 respectively. Triamcinolone showed affinity to NSP1 (**Figure 2**) and S protein with energies of −0.001145 and −0.00162, respectively. Amoxicillin showed affinity to viral proteins NSP1 and N with energies of −.0034 and −.0024, respectively. Hydrochlorothiazide showed affinity to Spike and NSP1 proteins with energies of −0.0057 and −0.0053 (**Table 2**). The energy units are in 0.0188 Hartrees.

**Figure 2:**
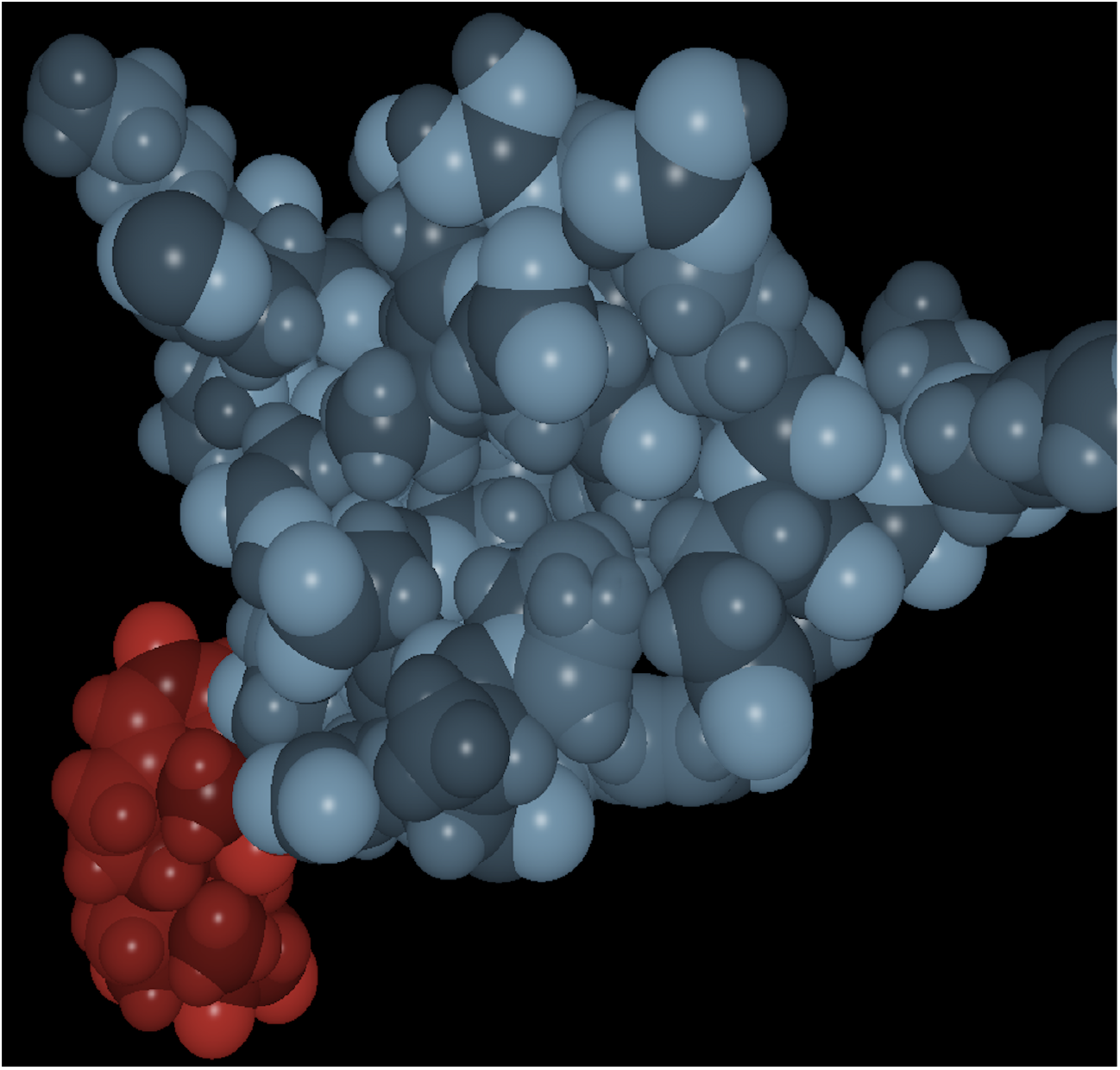
Image of triamcinolone (red) interacting with SARS-CoV-2 NSP1 protein (blue) at neutral pH. Darker shades signify regions of positive partial charges and lighter shades signify regions of negative partial charges.

*Diabetic (30-85yo, CCI<=3) and separately non-diabetic patients (30-85yo, CCI <= 3)* Results for ‘only diabetic patients’ showed Hydrochlorothiazide and Metformin had statistically significant reduction in mortality odds by 49% and 34%, respectively. Results for ‘only non-diabetic patients’ showed Hydrochlorothiazide and Metformin had statistically significant reduction in mortality odds by 30% each (**Table 3**).

## DISCUSSION

We identified four FDA-approved drugs as COVID-19 repositioning candidates using our novel in-silico quasi-quantum simulation methods followed by statistical analysis of 1.5M patients’ EHR data. We found that Metformin, Triamcinolone, Amoxicillin and Hydrochlorothiazide were associated with 27%, 26%, 26%, and 23% reduced mortality odds, respectively.

We highlight that among 1,513 drugs used in our in-silico simulations against specified 11 SARS-CoV-2 proteins, Metformin had the strongest in-silico signal (interaction energy − 0.007279) and also the highest reduction (by 27%) in COVID-19 mortality odds. It was followed by Hydrochlorothiazide’s signal (interaction energy of −0.005759) and 23% reduction in mortality odds.

The identified drugs Metformin (anti-diabetic), Triamcinolone (anti-inflammatory), Amoxicillin (anti-bacterial) and Hydrochlorothiazide (anti-hypertensive) are not members of a single class of drugs. Rather, each of these drugs have unique known clinical effect mechanisms, which may have played crucial roles in addition to their in-silico predicted SARS-CoV-2 protein interaction effect - both contributing to improved COVID-19 outcomes. The precise mechanisms by which these drugs exert their positive effects in COVID-19 will require further investigation and are beyond the scope of this study. For example, Metformin (a) inhibits gluconeogenesis thus reducing blood sugar^27^ and (b) helps activate pro-survival kinase AMPK which via mitochondria involved metabolic pathways results in cardiovascular health and lifespan improvement.^28^ Triamcinolone, a synthetic glucocorticoid, is used to treat autoimmune diseases, asthma, rheumatoid and arthritic conditions.^29,30,31^ The beta-lactam antibiotic, Amoxicillin, inhibits transpeptidation required for bacterial cell membrane synthesis.^32^ The loop diuretic, Hydrochlorothiazide, inhibits distal convoluted tubule renal sodium chloride transporter resulting in the loss of sodium and potassium, and reduction in blood pressure.^33^ Other researchers have also hypothesized Metformin’s positive effect on COVID-19.^34^

The identified drug candidates are usually well tolerated but caution must be followed for comorbid patients. Metformin, which has a 90% clearance via kidney tubular mechanism35, is contraindicated in patients with decreased creatinine clearance (CrCl) (< 45 ml/min) due to lactic acidosis risk.^36^ Amoxicillin is contraindicated for those with severe penicillin allergies. A CrCl based dose adjustment is necessary for ideal candidates with kidney disease. High risk patients with radiographically proven covid-19 pneumonia are selectively treated with antibiotics for possible superimposed bacterial infections with macrolides, cephalosporins or fluoroquinolones. A dose adjusted switch to amoxicillin for lower respiratory tract infection to treat community acquired pneumonia is feasible. Hydrochlorothiazide (HCTZ) is a diuretic used as a first line antihypertensive for essential hypertension patients. HCTZ can cause rare organ threatening complications like pancreatitis. Loop diuretics like Furosemide are being used in critically ill patients to maintain a negative fluid balance. Although not as potent as loop diuretics, adding HCTZ in select patients based on CrCl may prove to be beneficial to achieve diuresis37 and antiviral effect.

Triamcinolone can be administered systemically, orally, or by nebulization for direct pulmonary delivery. Thus it may act through both (a) interaction with SARS-CoV-2 proteins, and (b) pulmonary anti-inflammatory effects by stabilizing mast cells, a major cytokine storm source in COVID-19.^31^

Whether the doses and administration routes of these four drugs affect the primary outcome require further studies. Understanding the mechanisms by which these four drugs improve COVID-19 outcomes but not other drugs with similar functions (e.g. Fluticasone, or Clindamycin) will require further studies.

### Limitations

The N3C dataset did not track whether a patient was involved in a COVID-19 vaccination trial which, while unlikely, may skew results as vaccinated individuals are less likely to die from COVID-19. Our statistical procedure’s uncertainty intervals did not take into account the selection procedure for the propensity model nor for the implicit multiple comparison post estimation. Given multiple treatments of interest, and varying sample sizes for each treatment, accounting for these factors is nontrivial and we are not aware of any currently available method to accurately account for them. This could potentially lead to optimistic uncertainty estimates potentially inflating the type I error. Finally, a patient’s diabetic disposition was solely based on clinical diagnosis and did not take HbA1c levels into consideration to compare between controlled versus uncontrolled diabetes.

### Future work

Depending on funding we may look at the effect of these compounds on hypertensive and hospitalized patient subsets in addition to *in vitro* and *in vivo* antiviral assays. The novel simulation platform and the methodology to assess clinical effects we used have implications much beyond SARS-CoV-2.

## Data Availability

National Institute of Health (NIH) National COVID Cohort Collaborative (N3C) has clear procedures for researchers to gain access to the de-identified patient data (1000+ researchers already have access to the data)

https://www.ariscience.org/p1_sc2_paper_01.html

https://ncats.nih.gov/n3c

## ACKNOWLEDGEMENTS

We thank Massachusetts Green High Performance Computing Center, University of Maine System Advanced Computing Group, University of North Dakota Computational Research Center, Intel Corporation and ARI Internal HPC group for computing power contribution. The statistical analyses we performed were conducted with data or tools accessed through the NCATS N3C Data Enclave and supported by NCATS U24 TR002306. This research was possible because of the patients whose information is included within the data and the organizations and scientists who have contributed to the on-going development of this community resource.^12^ This work also used resources via the COVID-19 HPC Consortium, a private-public effort to bring together government, industry, and academia who are volunteering free COVID-19 research compute-time.^39^ ARIScience funded this project.

We also acknowledge reference data compilation assistance by Randa Bdair, Jeff Butler, Ievgen Duboriz and Haven López.

## CONFLICTS OF INTEREST

Joy Alamgir is founder of ARIScience. Melissa Haendel is a co-founder of Pryzm Health.

**Supplemental Appendix** is available at: https://www.ariscience.org/p1_sc2_paper_01.html

## Notes

### Author Declarations

This Data Utilization request of this project (RP-392493) was approved by National Institute of Health (NIH) National COVID Cohort Collaborative (N3C) DUR Approval Committee.

## REFERENCES

1. CDC COVID Data Tracker. Centers for Disease Control and Prevention website. Updated March 12, 2021. Accessed March 13, 2021. https://covid.cdc.gov/covid-data-tracker/

2. COVID-19 Vaccines. Food and Drug Administration. Updated March 12, 2021. Accessed March 13, 2021. https://www.fda.gov/emergency-preparedness-and-response/coronavirus-disease-2019-covid-19/covid-19-vaccines

3. Corona Virus Disease 2019. Centers of Disease Control and Prevention website. Updated December 3, 2020. Accessed December 4, 2020. https://www.cdc.gov/coronavirus/2019-ncov/vaccines/8-things.html

4. Paltiel AD, Schwartz JL, Zheng A, Walensky RP. Clinical Outcomes Of A COVID-19 Vaccine: Implementation Over Efficacy. Health Affairs. 2020. DOI:10.1377/hlthaff.2020.02054

5. Thigpen CL, Funk C. Most Americans expect a COVID-19 vaccine within a year; 72% say they would get vaccinated. Updated May 21, 2020. Accessed December 9, 2020. https://www.pewresearch.org/fact-tank/2020/05/21/most-americans-expect-a-covid-19-vaccine-within-a-year-72-say-they-would-get-vaccinated/

6. Tyson, A, Johnson, C, Funk, C. US public now divided over whether to get COVID-19 vaccine. Updated September 17 2020. Accessed December 9, 2020. https://www.pewresearch.org/science/2020/09/17/u-s-public-now-divided-over-whether-to-get-covid-19-vaccine/

7. Funk C, Tyson, A. Intent to get a COVID-19 Vaccine Rises to 60% as Confidence in Research and Development Process Increases. Updated December 2, 2020. Accessed December 9, 2020. https://www.pewresearch.org/science/2020/12/03/intent-to-get-a-covid-19-vaccine-rises-to-60-as-confidence-in-research-and-development-process-increases/

8. McGinley L, Abutaleb Y, Johnson C. Pfizer tells US officials it cannot supply substantial additional vaccines until late June or July. Washington Post. Dec 8, 2020. Accessed December 9, 2020. https://www.washingtonpost.com/health/2020/12/07/pfizer-vaccine-doses-trump/

9. Kwok KO, Lai F, Wei WI, et.al. Herd immunity - estimating the level required to halt the COVID-19 epidemics in affected countries. J Infect 2020, Mar 21;80(6):E32–E33. DOI: 10.1016/j.jinf.2020.03.027

10. Pushpakom, S., Iorio, F., Eyers, P. et al.. Drug repurposing: progress, challenges and recommendations. Nat Rev Drug Discov 18, 41–58 (2019). DOI: 10.1038/nrd.2018.168

11. Jupyter. Project Jupyter. Updated December 10, 2020. Accessed December 11, 2020. https://jupyter.org

12. Melissa A Haendel, Christopher G Chute, Tellen D Bennett, et al. The National COVID Cohort Collaborative (N3C): Rationale, design, infrastructure, and deployment, JAMIA Aug 2020. DOI: 10.1093/jamia/ocaa196

13. CommonDataModel. Github https://github.com/OHDSI/CommonDataModel. Updated December 10, 2020. Accessed December 11 2020.

14. Parks JM, Smith JC. How to discover antiviral drugs quickly. NEJM. 2020 May 20. DOI: 10.1056/NEJMcibr2007042

15. Fontes R, Ribeiro JM, Sillero A. Inhibition and activation of enzymes. The effect of a modifier on the reaction rate and on kinetic parameters. Acta Biochim Pol. 2000;47(1):233-57. PMID: 10961698.

16. OMOP/N3C Templates and Codes. ARIScience. Updated December 18, 2020. Accessed December 19, 2020. https://www.ariscience.org/y1_omop_n3c.html.

17. Ho D, Imai K, King G, Stuart E. MatchIt: Nonparametric Preprocessing for Parametric Causal Inference. J. Stat. Softw. 2011;Vol. 42, No. 8, pp. 1-28. DOI: 10.18637/jss.v042.i08

18. Stuart E. Matching methods for causal inference: A review and a look forward. Stat Sci. 2010;25(1):1–21. DOI:10.1214/09-STS313

19. Goodrich B, Gabry J, Ali I, Brilleman S (2020). “rstanarm: Bayesian applied regression modeling via Stan.” R package version 2.21.1. https://mc-stan.org/rstanarm

20. Johnson S, Speedie S, Simon G, Kumar V, Westra B. Quantifying the Effect of Data Quality on the Validity of an eMeasure. Appl Clin Inform. 2017;8(4):1012–1021. DOI:10.4338/ACI-2017-03-RA-0042

21. Huang, Y., Yang, C., Xu, Xf. et al.. Structural and functional properties of SARS-CoV-2 spike protein: potential antivirus drug development for COVID-19. Acta Pharmacol Sin 41, 1141–1149 (2020). DOI: 10.1038/s41401-020-0485-4

22. Zeng W, Liu G, Ma H, et al. Biochemical characterization of SARS-CoV-2 nucleocapsid protein. Biochem Biophys Res Commun. 2020;527(3):618–623. DOI:10.1016/j.bbrc.2020.04.136

23. Hou Y, Okuda K, Edwards C, et al. SARS-CoV-2 Reverse Genetics Reveals a Variable Infection Gradient in the Respiratory Tract. Cell. 2020;182(2):429-446.e14. DOI:10.1016/j.cell.2020.05.042

24. Schubert, K., Karousis, E.D., Jomaa, A. et al.. SARS-CoV-2 Nsp1 binds the ribosomal mRNA channel to inhibit translation. Nat Struct Mol Biol 27, 959–966 (2020). DOI: 10.1038/s41594-020-0511-8

25. O’Driscoll, M., Ribeiro Dos Santos, G., Wang, L. et al.. Age-specific mortality and immunity patterns of SARS-CoV-2. Nature (2020). DOI: 10.1038/s41586-020-2918-0

26. Charlson M, Szatrowski TP, Peterson J, Gold J. Validation of a combined comorbidity index. J Clin Epidemiol. 1994 Nov;47(11):1245–51. doi: 10.1016/0895-4356(94)90129-5. PMID: 7722560.

27. Hundal R, Krssak M, Dufour S, et al. Mechanism by which metformin reduces glucose production in type 2 diabetes. Diabetes Dec 2000, 49 (12) 2063–2069; DOI: 10.2337/diabetes.49.12.2063

28. Ritsinger V, Lagerqvist B, Lundman P, et al. Diabetes, metformin and glucose lowering therapies after myocardial infarction: Insights from the SWEDEHEART registry. Diab Vasc Dis Res. 2020 Nov-Dec;17(6):1479164120973676. DOI: 10.1177/1479164120973676. PMID: 33231125

29. Fitzpatrick A, Szefler S, Mauger D, et al. Development and initial validation of the Asthma Severity Scoring System (ASSESS). J Allergy Clin Immunol. 2020 Jan;145(1):127–139. DOI: 10.1016/j.jaci.2019.09.018. Epub 2019 Oct 8. PMID: 31604088

30. Sidhu G, Preuss CV. Triamcinolone. 2020 Sep 28. In: StatPearls [Internet]. Treasure Island (FL): StatPearls Publishing; 2020 Jan. PMID: 31335029

31. Theoharides T, Conti P. Dexamethasone for COVID-19? Not so fast. J Biol Regul Homeost Agents. 2020 Jul-Aug,;34(3):1241–1243. DOI: 10.23812/20-EDITORIAL_1-5. PMID: 32551464

32. Huttner A, Bielicki J, Clements M, Frimodt-Møller N, Muller A, Paccaud J, Mouton J. Oral amoxicillin and amoxicillin-clavulanic acid: properties, indications and usage. Clin Microbiol Infect. 2020 Jul;26(7):871–879. DOI: 10.1016/j.cmi.2019.11.028. PMID: 31811919

33. Roush G, Sica D. Diuretics for Hypertension: A Review and Update. Am J Hypertens. 2016 Oct;29(10):1130–7. DOI: 10.1093/ajh/hpw030. PMID: 27048970

34. Bramante C. Outpatient Metformin Use for Covid-19. Updated November 27, 2020. Accessed December 15 2020. https://clinicaltrials.gov/ct2/show/NCT04510194

35. Somogyi A, Stockley C, Keal J, Rolan P, Bochner F. Reduction of metformin renal tubular secretion by cimetidine in man. Br J Clin Pharmacol. 1987 May;23(5):545–51. DOI: 10.1111/j.1365-2125.1987.tb03090.x.

36. Salpeter SR, Greyber E, Pasternak GA, Salpeter EE. Risk of fatal and nonfatal lactic acidosis with metformin use in type 2 diabetes mellitus. Cochrane Database Syst Rev. 2010 Apr 14;2010(4):CD002967. doi: 10.1002/14651858.CD002967.pub4.

37. Magrina N, Marata A, Mario S, Daya L, Vignatelli L. Review of the available evidence on Thiazides Diuretics in the management of Heart Failure. WHO. Mar 2009. https://www.who.int/selection_medicines/committees/expert/17/application/thiazides.pdf

38. A Review of the SARS-CoV-2 (COVID-19) Genome and Proteome. Genetex. Updated April 21, 2020. Accessed December 3, 2020. https://www.genetex.com/MarketingMaterial/Index/SARS-CoV-2_Genome_and_Proteome

39. COVID-19 HPC Consortium. Updated December 14, 2020. Accessed December 14, 2020. https://covid19-hpc-consortium.org.

